# Safety and efficacy of COVID-19 hyperimmune globulin (HIG) solution in the treatment of active COVID-19 infection- Findings from a Prospective, Randomized, Controlled, Multi-Centric Trial

**DOI:** 10.1101/2021.07.26.21261119

**Authors:** Devang Parikh, Alok Chaturvedi, Naman Shah, Piyush Patel, Ronak Patel, Suma Ray

## Abstract

**Background:** COVID-19 hyper-immune globulin (HIG) solution is a human plasma-derived, highly-purified, concentrated, virus-inactivated preparation of neutralizing antibodies (NAbs) against COVID-19.

**Methods:** This was a randomized, two-arm, controlled, multi-center trial to evaluate the efficacy and safety of COVID-19 HIG in patients who were hospitalized with moderate-severe COVID-19 infection.

**Results:** A total of 60 patients were randomized (30 in each arm). Overall, COVID-19 HIG was well-tolerated without any serious treatment-emergent adverse event or tolerability issue. The mean change in ordinal scale by day 8 was 1.7±1.61 in the test arm vs. 2.0±1.68 in the control arm (mITT; p=0.367). Early and high NAbs were observed in the test arm compared to the control arm.

More patients had negative RT-PCR by day 3 for the test arm vs. the control arm (mITT: 46.67% in test vs. 37.93% in control). The median time to be RT-PCR negative was 5.5 days for the test arm vs. 8.0 days for the control arm for PP population. Patients receiving COVID-19 HIG showed early improvement (reduction) in the biomarkers (CRP, IL-6, and D-dimer).

**Conclusion:** COVID-19 HIG was found to be safe and well-tolerated. Early and high NAbs were achieved in COVID-19 HIG recipients qualifying the product as a suitable treatment option, particularly in an immunocompromised state. It should be given early in infection to mitigate progression to severe disease. It should be evaluated for post-exposure prophylaxis as well as for prevention (where a vaccine is not suitable or effective). It should be evaluated in the pediatric population as well.

## Introduction

Hyperimmune globulin is prepared from the human plasma with high titers of antibodies against a specific antigen. Some organisms against which hyperimmune globulins are available include hepatitis B, rabies, tetanus toxin, varicella-zoster, etc.^1^ Administration of hyperimmune globulin provides neutralization specific for diseases such as toxins, viruses, or bacteria.

A novel coronavirus disease (COVID-19) caused by the severe acute respiratory syndrome coronavirus 2 (SARS-CoV-2) has aroused an outbreak worldwide causing more than 3.3 million deaths.^2^ SARS-CoV-2 is highly infectious where most individuals are either asymptomatic or have mild-to-moderate symptoms. However, a substantial proportion has a severe, life-threatening disease course associated with a deleterious host immune response phase.^3^

In the absence of established therapeutic options for COVID-19, as a rapid response to the pandemic, convalescent plasma collected from the recovered patients was considered for the treatment to offer passive immunity to the patients.^4^ However, the plasma must be collected and processed from convalescent participants and verified to have adequate neutralizing antibodies. Apart from the anti-viral effects of the neutralizing antibodies in the plasma, it may exert an immunomodulatory effect in the hyperinflammatory state and competitively bind the Fc_γ_ receptor to prevent antibody-dependent enhancement triggered by virus-antibody immune complexes.^5^

Intravenous immunoglobulin (IVIG) infusion is associated with anti-inflammatory responses, including those from viral infection. A randomized placebo-controlled double-blind clinical trial in 59 patients with severe COVID-19 infection showed that administration of IVIG could improve their clinical outcome and significantly reduce the mortality rate.^6^ IVIG enriched with SARS-CoV-2 neutralizing antibodies have also been developed and being investigated for the treatment of COVID-19 infection.^7^ Clear demonstration of therapeutic benefit will require well-controlled studies.^8^

The use of hyperimmune globulin has proved clear efficacy in the treatment of influenza and SARS-CoV.^9^ Infusion of convalescent plasma to COVID-19 patients suggests the potential of hyperimmune globulin as a treatment to halt the progression of infection to severe pulmonary disease. Hyperimmune globulin treatment, besides inhibiting viral replication, can also down-regulate pro-inflammatory responses and can reduce disease severity in Covid-19 patients.

High dose antibodies could bind several different inhibitory receptors, including the inhibitory Fc gamma receptor or other receptors to induce the anti-inflammatory response. Regardless of the mechanism, the presence of neutralizing antibodies against SARS-CoV-2 as well as a high concentration of total antibodies could produce anti-inflammatory rather than the postulated immunopathology enhancement effects in COVID-19 participants to improve the prognosis.^10^

Globally, hyperimmune globulins prepared from the convalescent plasma are under clinical development for the prevention and treatment of COVID-19 infection.^11,12^ Additionally, recombinant monoclonal antibodies have been developed by some of the manufacturers. Casirivimab and Imdevimab have been given an emergency use authorization (EUA) by the US Food and Drugs Administration (FDA) for the treatment of mild to moderate COVID-19 in adults and pediatric patients.^13^ The antibody cocktail was also recently approved by the regulators in India.^14^

### Rationale and Aim

COVID-19 is a respiratory disease caused by a novel coronavirus (SARS-CoV-2) and causes substantial morbidity and mortality. There are very limited therapeutic agents for the management of COVID-19 infection and long-term efficacy and safety data of the vaccines are still awaited. This clinical trial was designed to evaluate the COVID-19 Hyper-Immune globulin (HIG) solution developed by Intas Pharmaceuticals Limited, India for the treatment of patients with active COVID-19.

The primary objective was to compare the efficacy of treatment with COVID-19 HIG plus standard of care versus only standard of care in participants with active COVID-19. Secondary objectives comprised of various efficacy parameters, neutralizing antibodies, and safety with COVID-19 HIG plus standard of care versus only standard of care in participants with active COVID-19.

## Methods

This was a prospective, open-label, two-arm, randomized, controlled, multi-centric trial for evaluation of efficacy and safety of COVID-19 HIG solution manufactured by Intas Pharmaceuticals Ltd. India. The trial was conducted in accordance with the applicable ethical standards and Good Clinical Practices guidance at seven sites after obtaining ethics committee approval and registration on the clinical trial registry of India (CTRI/2020/09/027903). All the trial participants had signed the informed consent document.

There were 2 days of screening period followed by 28 days of the study period. Participants were randomized in a 1:1 ratio in the test arm (T) and control arm (R). Key eligibility criteria were age between 18 to 65 years, laboratory-confirmed SARS-CoV-2 infection as determined by reverse transcription-polymerase chain reaction (RT-PCR) within <72 hours before randomization, moderate or severe active COVID-19 (Clinical Management of COVID-19 Guidelines of MOHFW Ver. 05), the female participant having a negative pregnancy test. Participants were excluded who had more than 5 days of COVID-19 specific hospitalization, had symptoms for more than 10 days, required invasive ventilation, or having hemodynamic instability (MOHFW guideline) or multiple organ dysfunction/failure or evidence of bacterial super-infection. Detailed eligibility criteria have been provided in appendix I.

Patients in the test arm received COVID-19 HIG, a purified SARS-CoV-2 hyper-IgG preparation rich in NAbs against COVID-19.^15^ COVID-19 HIG (350 AU/mL) was given as 30 mL intravenous infusion on Day 1 & 2 (at the same time preferably) plus standard of care as defined in guidelines on clinical management of COVID-19 issued by the Ministry of Health and Family Welfare, Government of India. Patients in the control arm received only standard of care.

### Statistical analysis

Descriptive analysis was done by tabulation of data and presentation of continuous variables as means and standard deviations or medians and ranges, as appropriate, and categorical variables as proportions. Baseline and demographic characteristics were summarized using descriptive statistics by study treatments.

For the primary outcome of the mean change from day 1 to day 8 in an 8-point ordinal scale, descriptive statistics were calculated and compared using the Wilcoxon rank-sum test. Secondary endpoints were analyzed as per the protocol. We compared changes in continuous variables such as oxygen requirement, laboratory parameters (biomarker levels, neutralizing antibody titers) throughout hospital stay between both arms by using generalized estimating equations. To assess viral clearance, we compared the proportion of participants negative for SARS-CoV-2 RNA between the trial arms on days 3 and 7, and 14.

Descriptive statistics were reported for time to clinical improvement (TTCI) defined as the time (in days) from initiation of study treatment until an improvement of two categories from enrolment status by day 14 on an 8-point ordinal scale of clinical status and compared using Wilcoxon rank-sum test. Descriptive statistics were to be reported for time to resolution of symptoms (shortness of breath, fatigue, cough, and fever) based on a 5-point ordinal scale up to 14 days. Count and percentage were provided for all-cause mortality at day 28 and proportion of patients negative for SARS-CoV-2 by RT-PCR at Days 3, 8, and 14.

A modified intention-to-treat analysis was performed on a subgroup of participants who have received at least one dose of study intervention and had undergone at least one post-dose efficacy evaluation. A post hoc subgroup analysis was conducted for the patients with a history of diabetes mellitus for the primary endpoint.

## Results

Out of the 60 eligible patients randomized (30 each in test and control arm), 53 patients (26 in test and 27 in control arm) were considered for the per-protocol set (PP), and 59 patients (30 in test and 29 in control arm) were considered for modified intention to treat (mITT) set.

Both the groups were comparable for the baseline characteristics of the participants (Table *I*). The mean age of the patients was 52±10.1 years [53±9.1 in the test arm and 52±11.2 in the control arm]. Out of 60 patients, 16 (26.7%) were female [6 (20.0%) in the test arm and 10 (33.3%) in the control arm] and 44 (73.3%) were male [24 (80.0%) in the test arm and 20 (66.7%) in the control arm].

**Table I.**
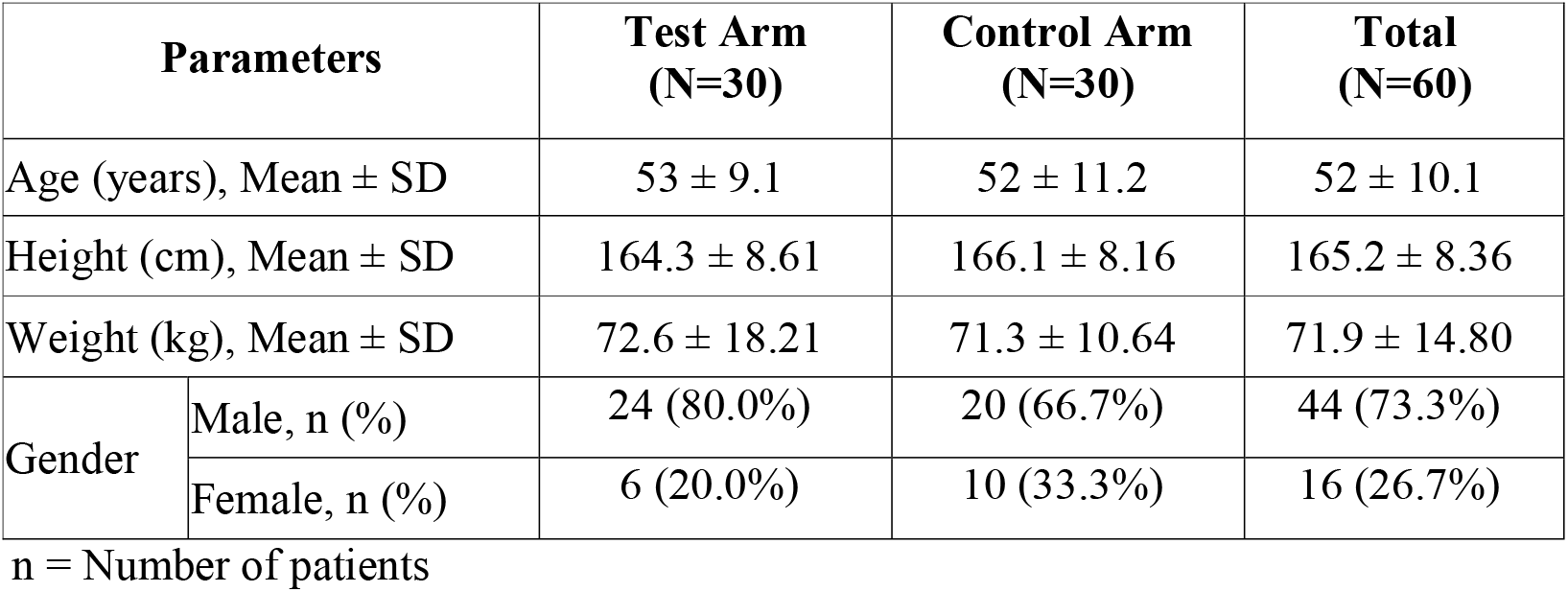
Baseline characteristics of the participants

### Efficacy

The mean change from baseline to day 8 in an 8-point ordinal scale in the mITT set was 1.7 ± 1.61 for the test arm vs. 2.0 ± 1.68 for the control arm.

Subgroup analysis in the patients with a history of diabetes showed that the mean change from baseline to day 8 in the ordinal scale was 1.8 ± 1.79 for the test arm vs. 1.3 ± 1.38 for the control arm (mITT set: 17 patients in test and 7 patients in control; p=0.77). The improvement in clinical outcome was comparatively better for the patients with diabetes mellitus who received COVID-19 HIG in addition to the standard of care.

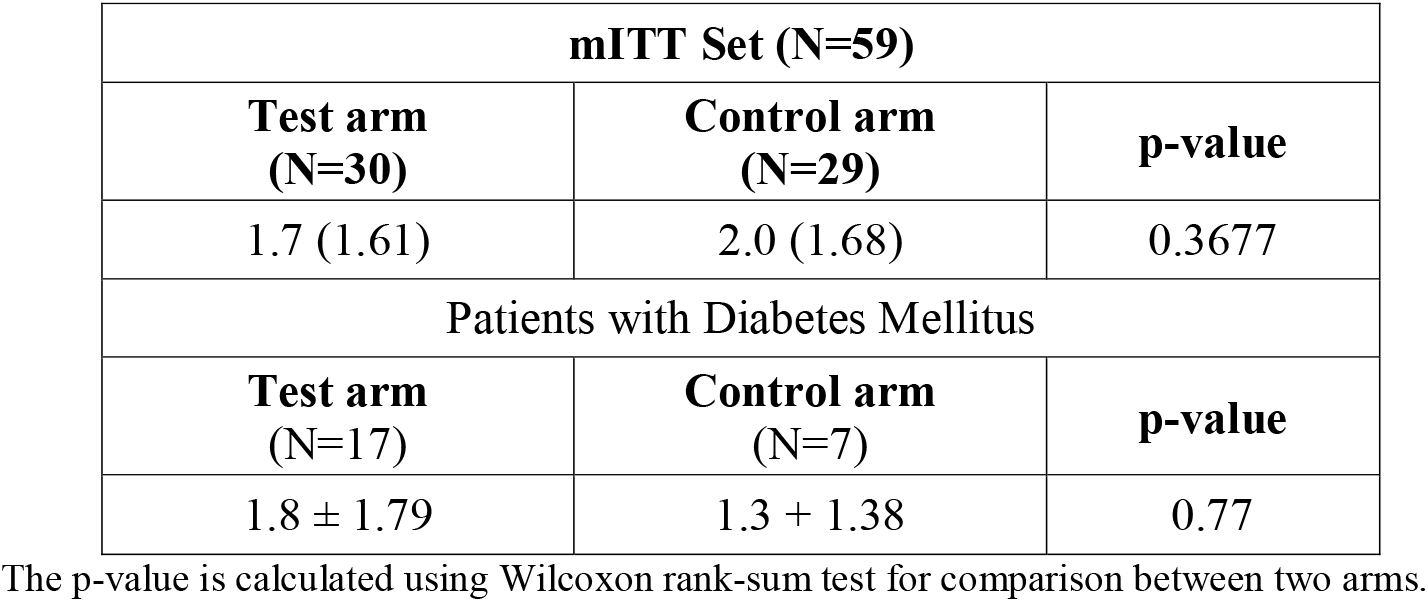

| mITT Set (N=59) |  |  |
| --- | --- | --- |
| Test arm<br>(N=30) | Control arm<br>(N=29) | p-value |
| 1.7 (1.61) | 2.0 (1.68) | 0.3677 |
| Patients with Diabetes Mellitus |  |  |
| Test arm<br>(N=17) | Control arm<br>(N=7) | p-value |
| $1.8 \pm 1.79$ | $1.3 \pm 1.38$ | 0.77 |

The p-value is calculated using Wilcoxon rank-sum test for comparison between two arms.

A relatively higher number of patients had early viral clearance as negative for SARS-CoV-2 by RT-PCR at day 3 after the treatment with COVID-19 HIG for mITT set. RT-PCR on day 3 was negative for 14/30 (46.67%) patients who received COVID-19 HIG along with the standard of care while it was negative for 11/29 (37.93%) patients who received only standard of care (Table *II*).

**Table II.**
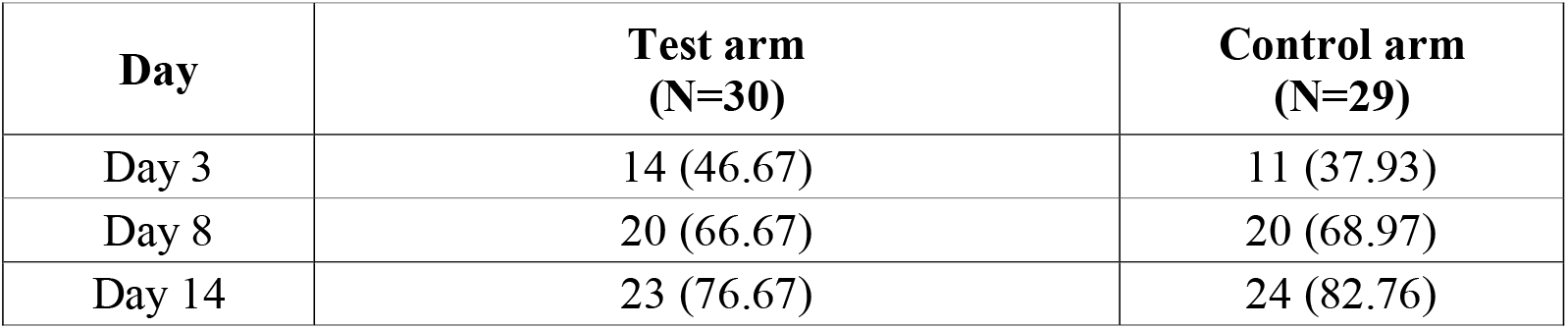
Count and percentage for negative SARS-CoV-2 by RT-PCR

The median time (in days) for RT-PCR to be negative was 5.5 days (range: 3 to 14 days) for the test arm while 8.0 days (range: 3 to 14 days) for the control arm (Figure *2*) in PP set. The median time to hospital discharge was 7.0 days for the test arm and 8.0 days for the control arm (mITT set). Only 10% of the test arm patients required hospitalization for more than 14 days versus 21% of control arm patients.

**Figure 1.**
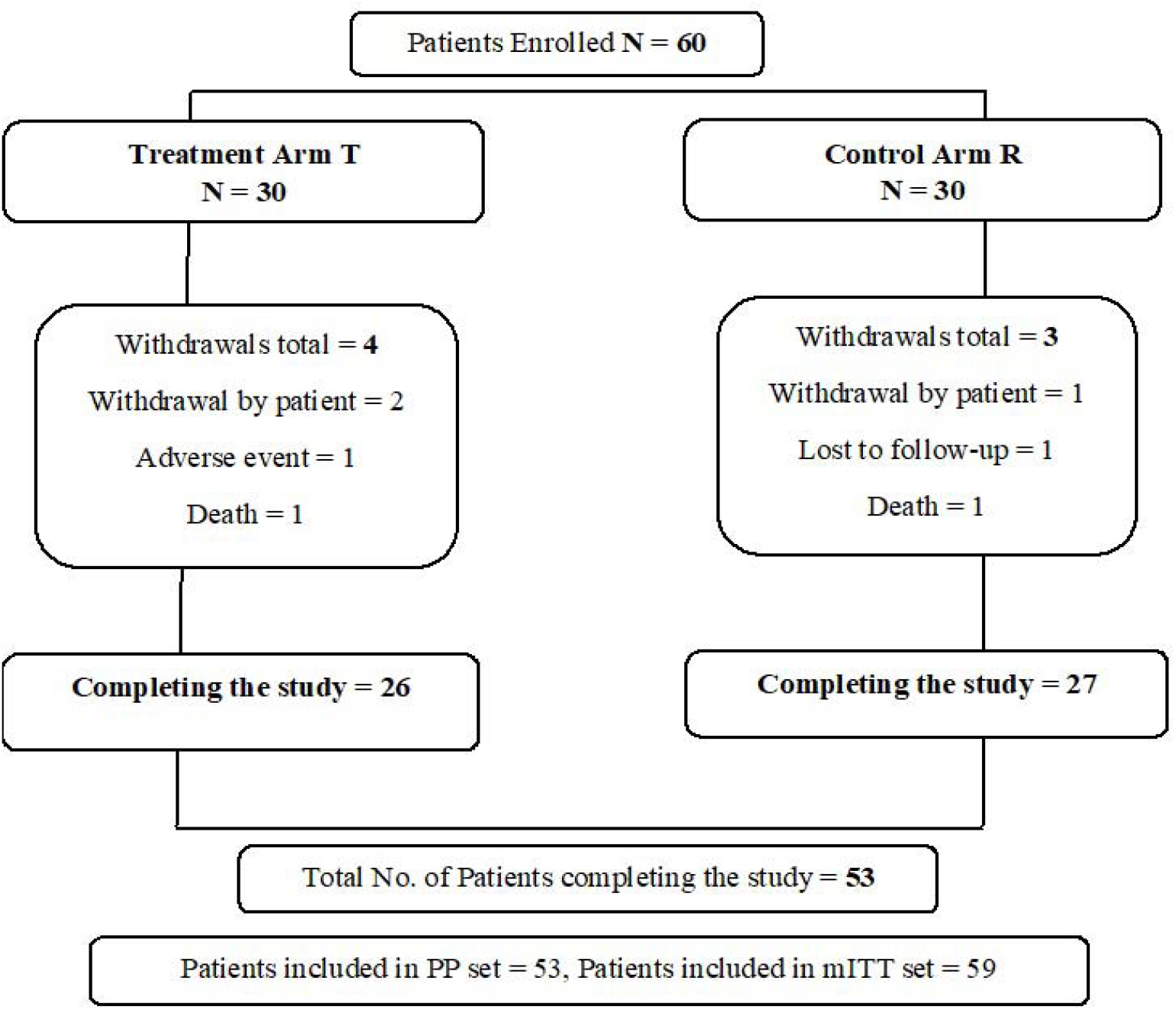
Patient disposition

**Figure 2.**
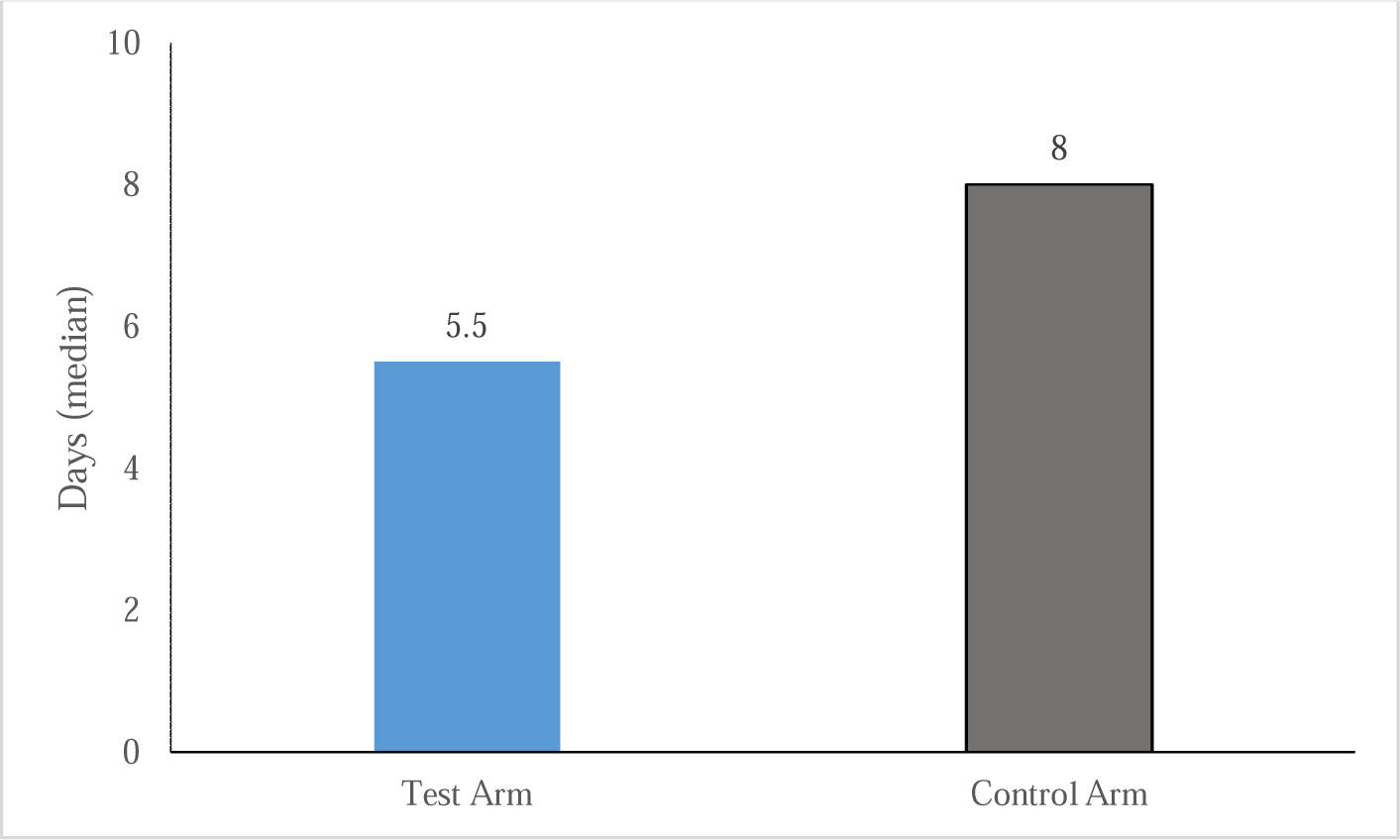
Median time to be RT-PCR negative (Days)

### Biomarkers evaluation

Biomarkers play a very important role in the prognosis of COVID-19 patients. The early and marked reduction was observed in the biomarkers following the COVID-19 HIG in comparison with the standard of care alone. Descriptive statistics for different biomarkers (CRP, IL6, D-dimer, and Ferritin) have been shown in Table *III* (mITT set).

**Table III.** Descriptive statistics for different biomarkers (CRP, IL6, D-dimer, and Ferritin) (mITT set)

| **Biomarkers** | **Time point** | **Test arm (N=30)**<br>**Mean (SD)** | **Control arm (N=29)**<br>**Mean (SD)** |
| --- | --- | --- | --- |
| **CRP**<br>**(mg/L)** | Day 1 (Baseline) | 83.1 (152.55) | 56.4 (53.55) |
|  | Day 3 | 33.5 (42.36) | 35.8 (59.63) |
|  | Day 8 | 11.2 (12.19) | 30.6 (105.20) |
|  | Day 14 | 11.6 (24.10) | 17.8 (40.97) |
| **IL6**<br>**(pg/mL)** | Day 1 (Baseline) | 37.3 (71.51) | 24.0 (25.15) |
|  | Day 3 | 20.4 (26.88) | 34.3 (82.14) |
|  | Day 8 | 16.3 (38.87) | 55.6 (201.49) |
|  | Day 14 | 44.1 (142.62) | 12.2 (11.93) |
| **D-dimer**<br>**(ng FEU/mL)** | Day 1 (Baseline) | 1282.2 (1865.37) | 807.6 (1331.73) |
|  | Day 3 | 1197.3 (1698.35) | 1095.6 (1620.24) |
|  | Day 8 | 949.7 (951.19) | 1087.9 (1609.35) |
|  | Day 14 | 722.3 (766.28) | 569.0 (449.15) |
| **Ferritin**<br>**(ng/mL)** | Day 1 (Baseline) | 484.6 (342.25) | 480.6 (424.99) |
|  | Day 3 | 462.1 (415.97) | 443.6 (340.64) |
|  | Day 8 | 417.7 (400.06) | 368.5 (422.33) |
|  | Day 14 | 350.3 (430.40) | 264.2 (207.11) |

The improvement of the clinical symptoms was comparable between the groups. Similarly, no major difference was observed in the supplemental oxygen requirements as assessed by the change in SpO2-FiO2 ratio.

### SARS-CoV-2 Neutralizing Antibodies (NAbs)

Early and high NAbs were observed in the test arm compared to the control arm. Patients who received COVID-19 HIG had sustained higher neutralizing antibody levels throughout the study duration (Figure 3). The NAbs on day 3 were three times higher than baseline for the patients who received COVID-19 HIG (percentage change from baseline in mean ± SD was 308.4±510.03 for the test arm and 186.7±331.10 for the control arm). Similarly on day 8 of the treatment, NAbs were raised approx. 7 times in the COVID-19 HIG recipients versus approx. 5 times in non-recipients with mean % difference of 253.3 between the arms (Table *IV*).

**Figure 3.**
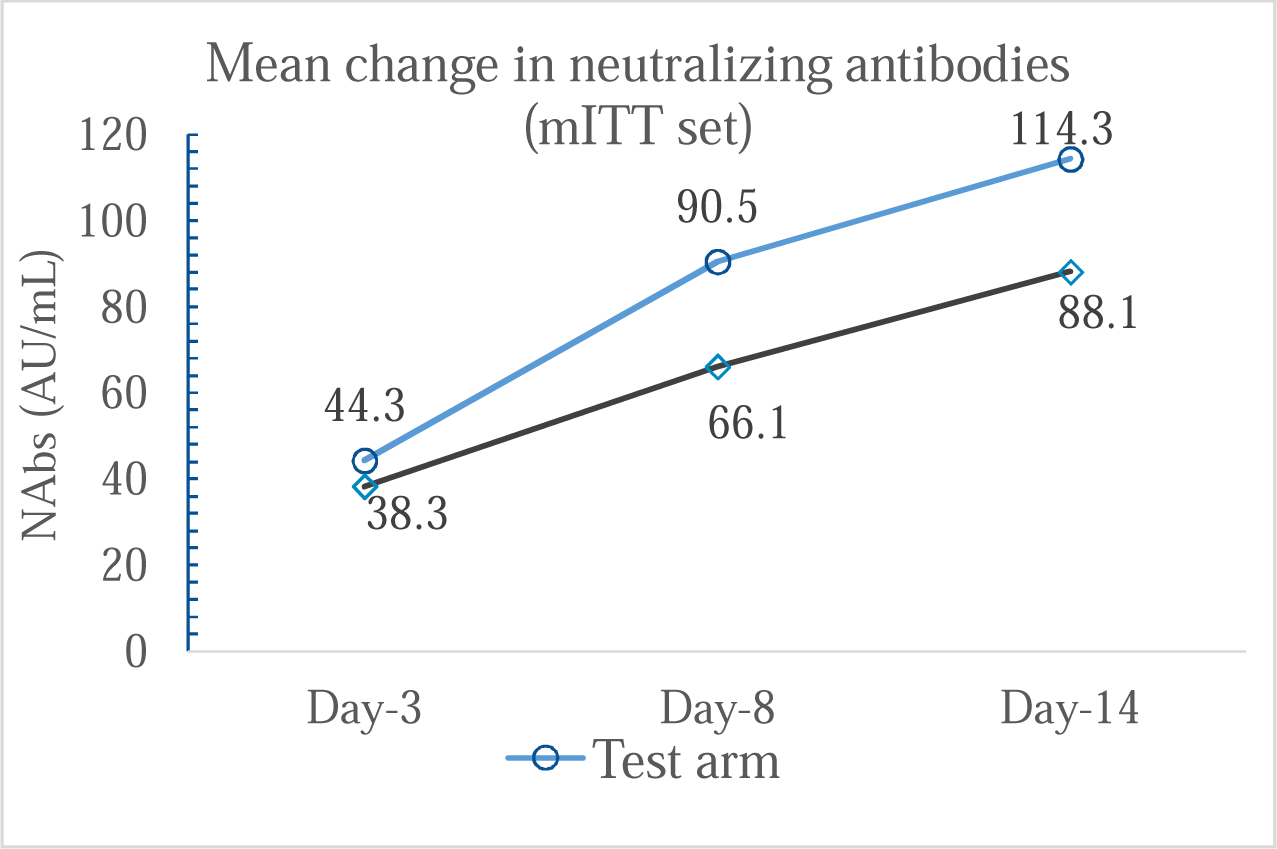
Mean change in neutralizing antibodies against SARS-CoV-2

**Table IV.** Percentage change in neutralizing antibodies from baseline (mITT set)

| Time point | Test arm<br>Mean (SD) | Control arm<br>Mean (SD) | Difference<br>in mean % |
| --- | --- | --- | --- |
| Day 3 | 308.4 (510.03) | 186.7 (331.10) | 121.7 |
| Day 8 | 714.9 (878.99) | 461.6 (621.96) | 253.3 |
| Day 14 | 943.1 (1151.36) | 706.0 (970.67) | 237.1 |

### Safety

A total of nine adverse events (AEs) were reported in five (8.3%) patients during the conduct of the study. Six AEs were reported in the test arm and 3 AEs were reported in the control arm (Table *V*). The causality assessment was judged as unlikely to the study intervention for 6 AEs and as possible for 2 AEs to the study drug administered. The causality assessment of the remaining 1 AE was not known.

**Table V.**
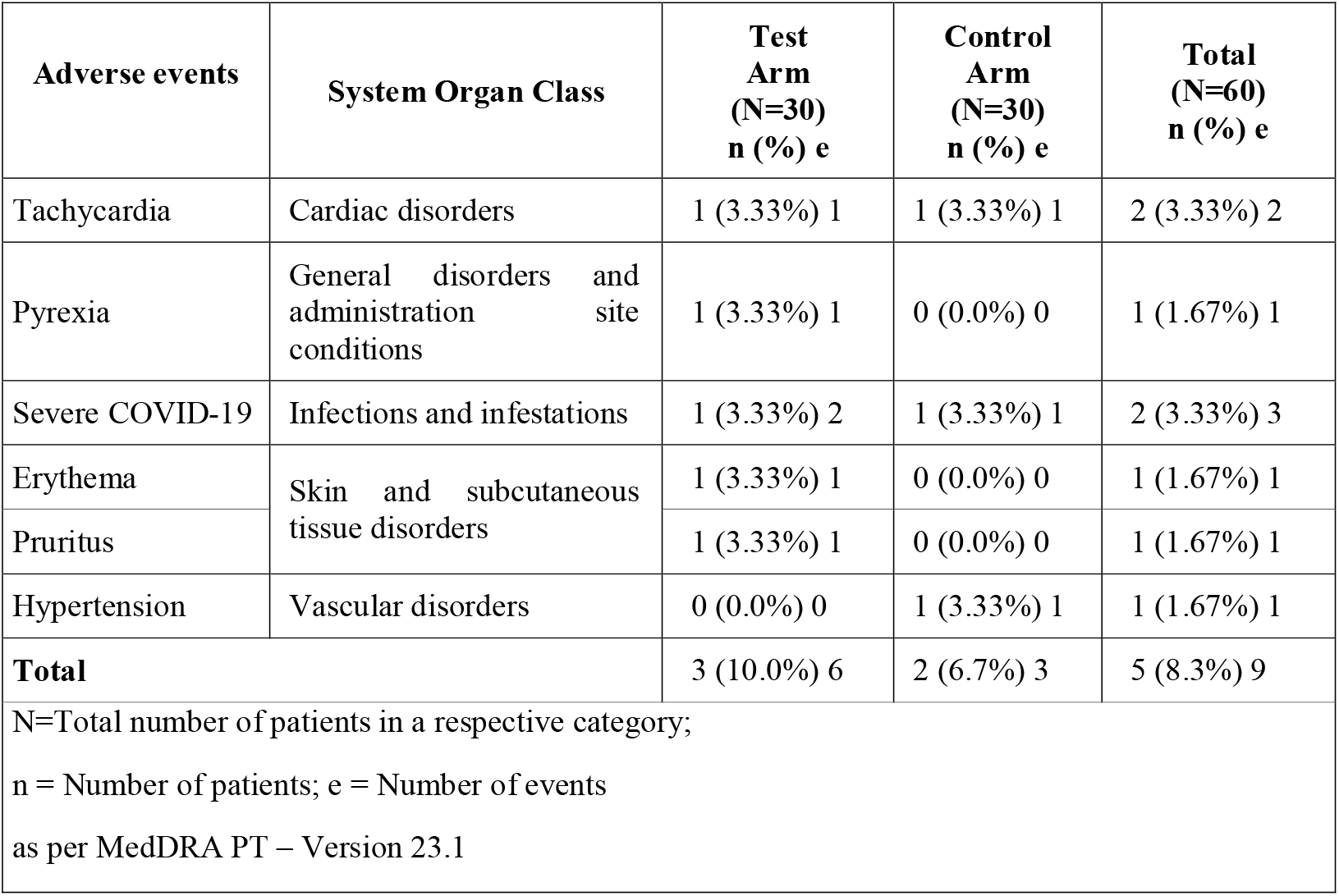
Adverse events

There were 2 serious adverse events (SAEs) (1 SAE in each arm) reported. The causality assessment was judged as unlikely for both the SAEs to the study drug administered. The event for both the SAEs was the severe COVID-19 and the outcome of both the SAEs was death. There were no other deaths or significant AEs during the conduct of the trial.

## Discussion

The COVID-19 pandemic has affected millions of lives globally.^16^ The evidence and understanding around the COVID-19 infection are still evolving. Two interconnected stages of the disease are generally recognized; an initial viral stage and a subsequent immune response phase with the clinical characteristics of hyper inflammation associated with acute respiratory distress syndrome. It has been proposed that the timing of the therapeutics is very important in the management of the COVID-19 disease.^17^ Therapies dosed soon after peak viral load when symptoms develop may decrease shedding duration and immune response intensity.^18^

The use of passive immunity using convalescent plasma or hyperimmune globulin is well established for many viral infections in the past.^19^ Convalescent plasma (CP) is a safe and potentially effective strategy for the treatment of emerging and re-emerging pathogens. The potential antiviral and immunomodulatory effects of CP have been explored for COVID-19 in clinical trials. It has been advised that the high-titer CP should be used as early as possible for the maximum benefit.^20^

A potential advantage of COVID-19 HIG over direct administration of single donor plasma or a monoclonal antibody therapy is the diversity of neutralizing antibodies obtained from a pool of convalescent donors.^21^ Neutralizing antibody diversity will cover the variants from the local circulation providing a broader range of anti-viral activity by attacking different viral epitopes thereby reducing the risk of promoting the mutations.

Intas manufactured COVID-19 HIG has been prepared from the convalescent plasma of the recovered patients having very high neutralizing antibodies against the SARS-CoV-2 virus.^15^ Purified and concentrated from pooled CP sourced from recovered patients and could be more beneficial than classical single-donor CP therapy. It can provide more consistent antibody levels in each lot, and units would be easy to store, distribute, and administer to patients. NAb titers observed in the serum microneutralization assay for final purified samples were >1:2,560 to <1:5,120 (1:640 for CP samples). The plaque reduction neutralization test (PRNT_90_) values in the final purified preparation were >1:640 (CP samples had >1:80 to <1:160).^22^

Another therapeutic regime introduced recently is the use of a single or cocktail of monoclonal antibodies. These are homogeneous and selective for a specific epitope of the virus which is not suitable for frequently changing RNA viruses.^23^ Furthermore, monoclonal antibodies may lead to a relatively higher chance of selection pressure to promote escape-mutation.^24^ Contrary to this, the polyclonal COVID-19 HIG is more versatile and has better coverage for virus strains.

In line with the expected characteristics of the plasma-derived products, COVID-19 HIG was safe and well-tolerated without any major safety concerns in moderate to severely ill COVID-19 patients. Patients who received HIG had early higher neutralizing antibodies in comparison with the control arm. Moreover, the higher NAbs were maintained throughout the study follow-up of 14 days in these patients.

It is learned that diabetes is an independent risk factor for higher morbidity and mortality in COVID-19.^25^ Both the groups had shown similar improvement in the 8-point ordinal scale while the high-risk diabetic patients who received COVID-19 HIG had shown relatively better improvement. There was no statistically significant difference observed in the mean change in the ordinal scale, a primary endpoint of the trial. The ordinal scale was calculated from the status of hospitalization and supplemental oxygen need. Practice for both of these was highly variable across the country during the active pandemic situation. Moreover, among the patients with diabetes, a relatively higher improvement was observed in the COVID-19 HIG recipients in comparison with the SOC alone.

It has been postulated that the early clearance of viruses will improve the outcome.^26^ During the trial, we have observed that the patients who received COVID-19 HIG along with standard care were RT-PCR negative by a median of 2.5 days earlier than the control arm. This would certainly provide clinical benefit by limiting the morbidity and complications.^27^ Moreover, a relatively shorter discharge time was observed in the patients receiving COVID-19 HIG means minimum utilization of hospital resources as well as fewer treatment expenses.

It has been observed that the patients in the test arm had numerically higher baseline values of CRP, IL-6, and D-dimer indicating a more intense immunological response. COVID-19 HIG treatment has provided quick and effective control of the anti-inflammatory stage as evident by a better reduction in all three biomarkers in comparison to the control arm. The early control of these biomarkers observed with the infusion of COVID-19 HIG might have played an important role in mitigating the possible risk of cytokine storm in the patients.

Our results are in line with the findings observed for similar products. Hyperimmune anti-COVID-19 IVIG (C-IVIG) developed by Dow University of Health Sciences has been tested for the treatment of severe and critical COVID-19 patients.^28^ Statistically significant improvement was observed in the clinical outcome as evident by the median Horowitz index in the patients who received C-IVIG. Mortality was decreased to 25% in the intervention groups in comparison with the 60% observed for the control arm. Additionally, reduction of biomarkers was also seen in intervention groups further supporting the possible immune-modulatory role of hyperimmune globulins in the management of COVID-19 infection. This is in line with the recent population-based real-world evidence where the strong inverse correlation between convalescent plasma use and mortality per admission was observed in the USA.^29^ The authors concluded that the retreat from convalescent plasma usage might have resulted in as many as 29,000 excess deaths from mid-November 2020 to February 2021 in the ISA. These findings further support the benefit of passive immunity-based therapeutic options in the treatment of COVID-19 infection.

While the product proved to be safe and well-tolerated, we must understand that the lack of clarity in efficacy could be due to the fact that the trial was planned during the early days of the active pandemic when the understanding of the disease, the available infrastructure, and expertise was limited and evolving. However, the hypothesis of the potential benefit of passive immunity is well established for the treatment of viral infections. Although we provided the required training, we have to permit reasonable flexibility amid the pandemic situation in the country.

Further, the standard of care offered to the patients has been continuously evolving. Considering the ethics of Good Clinical Practices, when there are no established treatments available, we allowed a wide range of potential therapeutic options at the discretion of the investigators including anti-virals, antimalarials, antibiotics, Remdesivir, corticosteroids, etc. The impact of such variation can not be ruled out. Additionally, this proof of concept trial was conducted in a relatively small group of 60 patients which may not provide reliable confidence for observed outcome.

## Conclusion

COVID-19 HIG was found to be safe and well-tolerated for patients with COVID-19 infection. Early viral clearance was observed in the patients who received COVID-19 HIG in addition to standard care as compared to standard care alone. Early limitation of infection can potentially improve the clinical outcome of high-risk patients. Early and high neutralizing antibodies against SARS-CoV-2 were achieved following the infusion of COVID-19 HIG qualifying the product as a suitable treatment option for the COVID-19 infection, particularly in patients with comorbidities and an immunocompromised state where natural antibodies are not developed. It should be given early in infection to mitigate progression to severe disease. Additionally, COVID-19 HIG should also be evaluated for post-exposure prophylaxis as well as for prevention (where a vaccine is not suitable or effective) of COVID-19 infection. Considering the established safety of plasma-derived products in children, COVID-19 HIG could also be evaluated in the pediatric population.

## Supporting information

Appendix I: Detailed eligibility criteria

## Data Availability

The authors confirm that the data supporting the findings of this study are available within the article and its supplementary material.

## Acknowledgment

We acknowledge the management of Intas Pharmaceuticals Ltd. and Lambda Therapeutics Research Ltd. for providing an opportunity to work on COVID-19 HIG in the fight against the pandemic. Lambda Therapeutics Research Ltd. was a contract research organization for this trial. The sponsor is thankful to all the trial investigators and site staff for their support. The sponsor is thankful to all trial participants for their contribution to the trial.

## Author’s contribution

All the authors were contributed to the conceptualization, study protocol development, and conduct of the clinical trial. AC and SR were involved in the overall trial management. DP, NS, PP, and RP were involved in the clinical operations, data analysis, and study report preparation. DP has preliminarily drafted the manuscript. SR has revised the manuscript critically for important intellectual content. All authors have thoroughly reviewed, amended and approved the final manuscript.

## Ethical statement

This study was conducted in accordance with the International Conference on Harmonisation Guidance for Good Clinical Practice guidelines and all applicable local regulatory requirements and laws. The study was approved by the Central Licensing Authority, Directorate General of Health Services, India. The approval was also obtained from the ethics committees of all the participating study sites: Care Institute of Medical Sciences, Ahmedabad, India; Ganesh Shankar Vidyarthi Memorial Medical College, Kanpur, India; Adichunchanagiri institute of medical Sciences, Mandya, India; Centre for Research SRV Hospital, Mumbai, India; Sri Ramachandra Institute of Higher Education & Research, Chennai, India; Government medical College, Nagpur, India; and Gujarat Medical Education & Research Society Gotri Medical College, Vadodara, India. Participants provided written informed consent before any study-related procedures were performed.

## References

1 Keller MA, Stiehm ER. Passive immunity in prevention and treatment of infectious diseases. Clin Microbiol Rev. 2000; 13(4): 602–614.

2 WHO Coronavirus (COVID-19) Dashboard. Available from: https://covid19.who.int/, accessed on May 16, 2021.

3 Salzberger B, Buder F, Lampl B. Epidemiology of SARS-CoV-2. Infection 2020. Available from: https://doi.org/10.1007/s15010-020-01531-3.

4 USFDA: Convalescent plasma EUA letter of authorization. FDA NEWS Release. Aug 2020.

5 Rojas M, Rodríguez Y, Monsalve DM, et al. Convalescent plasma in Covid-19: Possible mechanisms of action. Autoimmun Rev. 2020; 19(7): 102554.

6 Gharebaghi N, Nejadrahim R, Mousavi SJ. The use of intravenous immunoglobulin gamma for the treatment of severe coronavirus disease 2019: a randomized placebo-controlled double-blind clinical trial. BMC Infect Dis 2929; 20; 786.

7 Safety, PK and PD of Kamada Anti-SARS-CoV-2 in COVID-19. Last accessed on 3rd June 2021. Available from: https://clinicaltrials.gov/ct2/show/NCT04550325

8 Nguyen AA, Habiballah SB, Platt CD, Geha RS, Chou JS, McDonald DR. Immunoglobulins in the treatment of COVID-19: Proceed with caution!. Clinical Immunology. 2020; 11: 108459.

9 Arabi YM, Arifi AA, Balkhy HH, Najm H, Aldawood AS, Ghabashi A, et al. Clinical course and outcomes of critically ill patients with Middle East respiratory syndrome coronavirus infection. Ann Intern Med. 2014; 160(6): 389–97.

10 Ruklanthi de Alwis, Shiwei Chen, Esther S. Gan, Eng Eong Ooi, Impact of immune enhancement on Covid-19 polyclonal hyperimmune globulin therapy and vaccine development, EBio Medicine. 2020.

11 Vandeberg P, Cruz M, Diez JM, et al. Brief report: Production of anti-SARS-CoV-2 hyperimmune globulin from convalescent plasma. Transfusion. 2021: 1–5.

12 Ali S, Uddin SM, Ali A, Anjum F, Ali R, Shalim E, et al. Production of hyperimmune anti-SARS-CoV-2 intravenous immunoglobulin from pooled COVID-19 convalescent plasma. Immunotherapy. 2021; 13(5): 397–407.

13 U.S. Food and Drug Administration. Coronavirus (COVID-19) Update: FDA Authorizes Monoclonal Antibodies for Treatment of COVID-19. 2020. Available from: https://www.fda.gov/news-events/press-announcements/coronavirus-covid-19-update-fda-authorizes-monoclonal-antibodies-treatment-covid-19. accessed on June 8, 2021.

14 Cipla: Roche receives EUA India investigational antibody cocktail casirivimab Imdevimab covid. Available from: https://www.cipla.com/press-releases-statements/roche-receives-EUA-India-investigational-antibody-cocktail-casirivimab-Imdevimab-covid, accessed on June 8, 2021.

15 Verma S, Dolia S, Pawar A, Ray S. SARS-CoV-2 Hyper-Immunoglobulin: Purification and Characterization from Human Convalescent Plasma. BioProcess Int. 2021; 19(4).

16 Blake P. 2020 Year in Review: The impact of COVID-19 in 12 charts. World Bank Blogs. Dec 2020. Available from: https://blogs.worldbank.org/voices/2020-year-review-impact-covid-19-12-charts

17 Goyal A, Cardozo-Ojeda EF, Schiffer JT. Potency and timing of antiviral therapy as determinants of duration of SARS-CoV-2 shedding and intensity of inflammatory response. Science Advances 2020; 6(47): eabc7112.

18 S. Zheng, J. Fan, F. Yu, B. Feng, B. Lou, Q. Zou, G. Xie, et al. Viral load dynamics and disease severity in patients infected with SARS-CoV-2 in Zhejiang province, China, January-March 2020: Retrospective cohort study. BMJ 2020; 369: m1443.

19 Keller MA, Stiehm ER. Passive Immunity in Prevention and Treatment of Infectious Diseases. Clin Microbiol Rev. 2000; 13(4): 602–614.

20 Bratcher-Bowman N. USFDA: Convalescent plasma emergency authorization. FDA NEWS Release. Mar 2021. Available from https://www.fda.gov/media/141477/download, accessed on May 23, 2021.

21 Liu L, Wang P, Nair MS, et al. Potent neutralizing antibodies against multiple epitopes on SARS-CoV-2 spike. Nature 2020; 584: 450–6.

22 Data on file. Intas Pharmaceuticals Ltd. 2020.

23 B.P. da Costa C, Martins FJ, et al. COVID-19 and Hyperimmune sera: A feasible plan B to fight against coronavirus, Int Immunopharmacology 2021; 90: 107220.

24 Chen J, Gao K, Wang R, Wei G. Revealing the threat of emerging SARS-CoV-2 mutations to antibody therapies. BioRxiv preprint. 2021. doi: https://doi.org/10.1101/2021.04.12.439473

25 Barron E, Bakhai C, Kar P, Weaver A, Bradley D, et al. Associations of type 1 and type 2 diabetes with COVID-19-related mortality in England: a whole-population study. Lancet Diabetes Endocrinol 2020; 8: 813–22.

26 Xue J, Zheng J, Shang X, et al. Risk factors for prolonged viral clearance in adult patients with COVID-19 in Beijing, China: A prospective observational study. Int Immunopharmacol. 2020; 89(Pt A): 107031.

27 Fajnzylber, J., Regan, J., Coxen, K. et al. SARS-CoV-2 viral load is associated with increased disease severity and mortality. Nat Commun 2020; 11: 5493.

28 Ali S, et al. Hyperimmune anti-COVID-19 IVIG (C-IVIG) treatment in severe and critical COVID-19 patients: A phase I/II randomized control trial, EClinicalMedicine (2021), https://doi.org/10.1016/j.eclinm.2021.100926.

29 Casadevall A, Dragotakes Q, et al. Convalescent Plasma Use in the United States was inversely correlated with COVID-19 Mortality: Did Plasma Hesitancy cost lives? MedRxiv preprint. 2021. doi: https://doi.org/10.1101/2021.04.07.21255089

