## Appendix I: Detailed eligibility criteria for "Safety and efficacy of COVID-19 hyperimmune globulin (HIG) solution in the treatment of active COVID-19 infection- Findings from a Prospective, Randomized, Controlled, Multi-Centric Trial"

Detailed eligibility criteria for the trial participants:

Inclusion criteria:

1. Participant and/or legally acceptable representative must sign an ICF to participate in the study indicating that the participant understands the purpose of, and procedures required for the study as described in this protocol and is willing to and will be able to adhere to requirement of the protocol.
2. Participant must be 18 to 65 years of age (both inclusive), at the time of signing the informed consent.
3. Participants must have documented laboratory-confirmed SARS-CoV-2 infection as determined by reverse transcription- polymerase chain reaction (RT-PCR) in any specimen, within <72 hours prior to randomization.
4. Participants with moderate or severe active COVID-19 (Clinical Management of COVID-19 Guidelines of MOHFW) at screening and baseline defined as,
    a. Radiological evidence of pulmonary infiltrates or Clinical features such as dyspnea and/or hypoxia, fever, cough,
    b. $SpO_2$ of <94 % on room air, and
    c. Respiratory rate of ≥24 per minute.
5. A female participant is eligible to participate if she is not pregnant or breastfeeding, and at least one of the following conditions applies:
    a. Is not a woman of childbearing potential (WOCBP),

       Or

       Is a WOCBP and using an acceptable contraceptive method as described in Appendix 10.4 of the protocol during the intervention period and at a minimum 30 days until after the last dose of study intervention. The investigator should evaluate the effectiveness of the contraceptive method in relationship to the first dose of study intervention.
    b. A WOCBP must have a negative highly sensitive pregnancy test (serum) within 4 days before the first dose of study intervention.
    c. Additional requirements for pregnancy testing during and after study intervention are in Appendix 10.2 of the protocol.
    d. The investigator is responsible for review of medical history, menstrual history, and recent sexual activity to decrease the risk for inclusion of a woman with an early undetected pregnancy.
6. Male participants are eligible to participate if they agree to the following during the intervention period and for at least 90 days after the last dose of study intervention:
    a. Must agree not to donate sperm for the purpose of reproduction,

       PLUS
    b. Must agree to use contraception /barrier as detailed below:
        i. a male participant must wear a condom when engaging in any activity that allows for passage of ejaculate to another person.

ii. Should also be advised of the benefit for a female partner to use a highly effective method of contraception described in Appendix 10.4 of the protocol as a condom may break or leak when having sexual intercourse with a woman of childbearing potential who is not currently pregnant.

Exclusion criteria

1. Participant requiring invasive ventilation or having hemodynamic instability (MOHFW guideline) or multiple organ dysfunction/failure or evidence of bacterial superinfection (as defined by Procalcitonin level ≥0.5 µg/L or other applicable diagnostic parameters as per standard medical care) as per the independent clinical judgment of the Investigator at screening and /or baseline.
2. Documented medical history of known allergies, hypersensitivity, or intolerance to intravenous immunoglobulin or other injectable form of IgG or blood products.
3. Documented medical history of known IgA deficiency.
4. Participants with a lifetime history of at least one thrombotic event including deep vein thrombosis, cerebrovascular accident, pulmonary embolism, transient ischemic attacks, or myocardial infarction.
5. Participants who have received any blood products within 30 days prior to randomization.
6. Participant with more than 5 days of COVID-19 specific hospitalization prior to the first administration of treatment at baseline.
7. Participants who have more than 10 days between the onset of symptoms and the day of first administration of treatment at baseline.
8. Pregnant or breastfeeding female participants.
9. Currently receiving renal replacement therapy/dialysis OR Creatinine clearance < 50 mL/min using the Cockcroft-Gault formula.
10. Documented medical history of hepatitis B surface antigen (HbsAg) or hepatitis C antibody (anti-HCV) positive, or other clinically active liver disease, or tests positive for HbsAg or anti-HCV at Screening.
11. Documented medical history of human immunodeficiency virus (HIV) antibody positive, or tests positive for HIV at Screening.
12. Currently receiving or has received in the last 14 days, experimental immune modulators, and/or monoclonal antibody therapies.
13. Confirmed diagnosis of bacterial pneumonia or other active/uncontrolled fungal or viral infections at screening/baseline.
14. Participants who have received organ transplantation or major surgery in the past 6 months.
15. Participants whose ALT/AST levels are 5 times higher than the normal upper limit and total bilirubin is 3 times higher than the upper limit of normal.
16. Co-morbid systemic illnesses (uncontrolled diabetes, uncontrolled hypertension, cardiac disease, chronic lung disease, chronic kidney disease, immune-suppression and cancer or other severe concurrent disease) which, in

the judgment of the investigator, would make the participant inappropriate for entry into this study or interfere significantly with the proper assessment of safety and toxicity of the prescribed treatment.

17. Current participation in another interventional clinical trial (with an investigational drug) that is not an observational registry and have received an investigational intervention 30 days or 5 half-lives (whichever is longer) before the signing the consent.
18. Participation in any other clinical trial of an experimental treatment for COVID-19.
19. Any other clinical/social/ psychiatric condition for which, in the opinion of the investigator, participation would not be in the best interest of the participant (e.g. compromise the well-being) or that could prevent, limit, or confound the protocol-specified assessments.
20. Employee of the investigator or study site, with direct involvement in the proposed study or other studies under the direction of that investigator or study site.

All the enrolled patients fulfilled all the above inclusion and none of the exclusion criteria.
